# Agent-Based Simulation of Covid-19 Vaccination Policies in CovidSIMVL

**DOI:** 10.1101/2021.01.21.21250237

**Authors:** Ernie Chang, Kenneth A. Moselle

## Abstract

An agent-based infectious disease modeling tool (CovidSIMVL) is employed in this paper to explore outcomes associated with MRNA two-dose vaccination regimens set out in Emergency Use Authorization (EUA) documents submitted by Pfizer and Moderna to the US Department of Health & Human Services. As well, the paper explores outcomes associated with a third “Hybrid” policy that reflects ranges of expected levels of protection according to Pfizer and Moderna EUA’s, but entails a 35 day separation between first and second dose, which exceeds the 21 days set out in Pfizer documentation or the 28 days in Moderna documentation.

Four CovidSIMVL parameters are varied in the course of 75 simulated clinical trials. Two relate directly to the vaccines and their impacts (duration between doses; degree of expected protection conferred by different vaccines following first or second dose). Two relate to the simulation contexts to which the vaccines are applied (degree of infectivity; duration of infectivity). The simulated trials demonstrate expected effects for timing of second dose, and for degree of protection associated with first and second dose of Pfizer and Moderna vaccines, and the effects are consistent with an assumed value of 75% for degree of protection after first and second doses for the Hybrid vaccine. However, the simulated trials suggest a more complex interaction between expected level of protection following first dose, timing of second dose and degree of infectivity. These results suggest that policy options should not be considered independent of the transmission dynamics that are manifested in the contexts in which the policies could be applied.

CovidSIMVL embodies stochasticity in the mechanisms that govern viral transmission, and it treats the basic reproduction number (R0)as an emergent characteristic of transmission dynamics, not as a pre-set value that determines those dynamics. As such, results reported in this paper reflect outcomes that could happen, but do not necessarily reflect what is more or less likely to happen, given different configurations of parameters. The discussion section goes into some measure of detail regarding next steps that could be pursued to enhance the potential for agent-based models such as CovidSIMVL to inform exploration of possible vaccination policies, and to project outcomes that are possible or likely in local contexts, where stochasticity and heterogeneity of transmission must be featured in models that are intended to reflect local realism.

## BACKGROUND

This paper explores a method for evaluating vaccination policies *via* simulated clinical trials using CovidSIMVL ((github.com/ecsendmail/MultiverseContagion), an agent-based tool for simulating viral spread. ^1,2^ The method is illustrated with a set of simulated trials that are intended to supply information that can support efforts to optimize use of limited supplies of SARS-CoV-2 vaccines that require two doses to confer maximum protection (Pfizer^3^ at 21 days for the second dose after the first dose, Moderna^4^ at 28 days after the first dose). Specifically, two of three sets of trials reported in this document evaluate the impact a single dose (in order to maximize number of people with some protection) *vs* the full series of two doses, according to the schedules evaluated in Pfizer and Moderna stage 3 clinical trials.

The third set of trials evaluates a “Hybrid” approach that is abstracted and synthesized from a variety of published materials that are all concerned with reconciling the guidelines or expectations set out in Emergency Use Authorization applications from Pfizer or Moderna with vaccine supply requirements that outstrip vaccine supply and/or delivery systems for the current and reasonably foreseeable future. These materials include documents provided by the Canadian National Advisory Committee on Immunization^5^ (NACI), directives from the Public Health Officer of British Columbia,^6,7,8^ and the Strategic Advisory Group of Experts on Immunization (SAGE/World Health Organization).^9^

## MOTIVATION

The intent is to parameterize the space within which models of vaccination strategies should be configured, and to run an initial set of simulations incorporating those parameters, to gauge the potential for an agent-based model to provide visibility into possible outcomes by revealing the potential for changes in those parameters to affect outcomes. In the simulations reported in this paper, there are four factors varying in the trials:

1. Duration between first dose and second dose (3 values)
2. Degree of protection following first dose (4 values, including 0% protection for Baseline No Vaccine trials)
3. Infectivity (3 values)
4. Duration of infectivity, varying to simulate impact of contact tracing (2 values)

## CAVEATS

This is a methodological paper. Though it consists of a series of simulated clinical trials using an agent-based modeling tool to reflect different vaccine schedules, it is not intended to predict outcomes associated with any of those schedules. This limitation in intended scope is a reflection of the manner in which CovidSIMVL incorporates stochasticity (randomness) into transmission dynamics, in order to enable the tool to provide meaningful simulations of real-world localized scenarios that do not assume homogeneous transmission.

In all four factors as set out in the section entitled “Motivation” were incorporated into a fully crossed design, this would yield 72 different conditions. Due to stochasticity inherent in the transmission dynamics that determine reproduction rates in CovidSIMVL, each of these combinations of conditions would need to be run repeatedly to determine when and in what sense different outcomes could be treated as reflections of some sort of central tendency vs outliers. The trials reported in this paper (20 for Pfizer, 20 for Moderna, 25 for “Hybrid” and 10 for No Vaccinations) incorporate all of the different levels on all of these variables. However, the large number of repeat trials that would need to be carried

out to yield outcomes associated with each combination of parameters – and these outcomes would themselves need to be modeled – have not been run. As such, the results in this paper can be treated as an evaluation of the *viability* of the method but they cannot be treated as an evaluation of the *advisability* of any of the vaccination schedules embodied in the simulated clinical trials.

## SETTING VACCINATION-SCHEDULE-RELATED PARAMETERS FOR SIMULATIONS - REFERENCES

The three sets of simulations in this paper are organized around three reference authorities that are used to set timings for administration of two vaccines that have been approved for use in Canada as of January 14, 2021 (Pfizer, Moderna) and expected levels of protection if those timings are met.

- Documentation supplied by Pfizer in their Emergency Use Authorization requests (EUA) to the Vaccines and Related Biological Products Advisory Committee of the US Food and Drug Administration, a division of the US Department of Health & Human Service.
- Documentation supplied by Moderna in their EUA request to the Food and Drug Administration.
- For the Hybrid model – various sources including Canadian National Advisory Committee on Immunization (NACI), the BC Provincial Health Officer, SAGE/World Health Organization.

Pfizer specifies 21 days between doses, and Moderna specifies 28 days. NACI states that they could both be taken 28 days apart, though not interchangeably. BC Centre for Disease Control (BCCDC) and NACI estimate an efficacy level of 80% for Moderna starting at two weeks after first dose. There appears to be more diversity of thinking regarding Pfizer, with 54% appearing as a low-end estimate of protection after the first dose, but NACI suggesting it could be as high as 80%, in keeping with Moderna, given that both are MRNA vaccines. Various sources attribute different levels of confidence to different figures, depending on how long trials have run, and how many cases fit a particular condition.

In light of the above, the values set on parameters for the models as detailed below should be regarded as hypothetical. This is particularly true for the Hybrid model, where the values reflect several different sources.

## METHODS

### Vaccination Strategies

We simulate three strategies in this working paper:

#### Pfizer

1. Dose 1 at Time = 0 (T.0).
2. Protection of 54% at T>14 days.
3. Mode1 – Dose 2 at T.21 causes protection to go to 95% at T.22.
4. Mode2 – Dose 2 is not given, and protection goes to zero at T.22.

#### Moderna

1. Dose 1 at time Time = 0 (T.0).
2. Protection of 80% at T>14 days.
3. Mode1 – Dose 2 at T.28 causes protection to go to 95% at T.29.
4. Mode2 – Dose 2 is not given, and protection goes to zero at T.29.

#### Hybrid (35 days to second dose)

1. Dose 1 at Time = 0 (T.0).
2. Protection of 75% at T>14 days (“splitting the difference” between a low-end estimate of 54% and roughly 80%).
3. Mode1 – Dose 2 at T.35 causes protection to go to 95% at T.36.
4. Mode2 – Dose 2 is not given, and protection goes to zero at T.36.

In the material below, we refer to the no vaccination comparison condition as “**B**” (baseline).

### Vaccination Policies

The simulations are keyed to two candidate policy positions. The first is to hold back half the vaccine supply, to assure maximum protection associated with completion of the two-dose vaccination regimen. In the simulations, we refer to this as “Mode1”.

The second is to use all the vaccine, with no assurance that vaccine for a second dose with be forthcoming within a window of time required for optimal protection to be achieved. This produces a branch in the policy enactment, depending on supply. In the worst-case scenario, no supply is forthcoming on day 21/28/35 in which case we are looking at a single dose mode (“Mode2”).

Alternatively, if supplies enable a second dose to be administered in a timely fashion, the result is the first and best-case scenario.

### Assumption - abrupt transitions in vulnerability or protection

Presumably the vaccines do not function like “light-switches” at a person or population level, with protection suddenly appearing or disappearing over the course of very short timing intervals. For example, both Pfizer and Moderna indicate that protection builds from the time of the first dose to a reported level of protection at 14 days.

However, lacking data on gradients of protection, and to highlight the effects of different vaccine schedules and produce a clear set of initial results, we use the assumption that immunity starts at 14 days after first dose, for a period of 21 or 28 or 35 days, and then falls to zero on day 22 or 29 or 36, if the second dose is not administered. Based on available data, we assume 95% protection upon administration of the second dose for Pfizer, Moderna or Hybrid.

### Initialization of simulations

In the CovidSIMVL agent-based model, agents can be in any of the following states at any given point in time:

- Susceptible (red in CovidSIMVL visualizations)
- Exposed and incubating (yellow)
- Asymptomatic or undetected but infective (blue)
- Symptomatic (detected) and infective (red)
- Inert - recovered and assumed no longer infective; deceased; quarantined (orange)

See ***Figure 1*** for a screen shot of a CovidSIMVL simulation, with agents coloured according to the scheme set out directly above. Note that the outbreak depicted in this figure is one self-extinguishes before all Susceptibles have been infected, so there are only “greens” (Susceptibles) and “Oranges” (Inerts). There are no “Yellows” (incubating) or infectives (“Blues” or “Reds”). The same colour scheme is used for the portion in the upper right-hand quadrant of this simulation that depicts the history of the outbreak, with a decreasing number of Susceptibles, a peak of infectives (Red) in the middle, and an increasing and then leveling off number of Inerts as the outbreak self-limits

**Figure 1.**
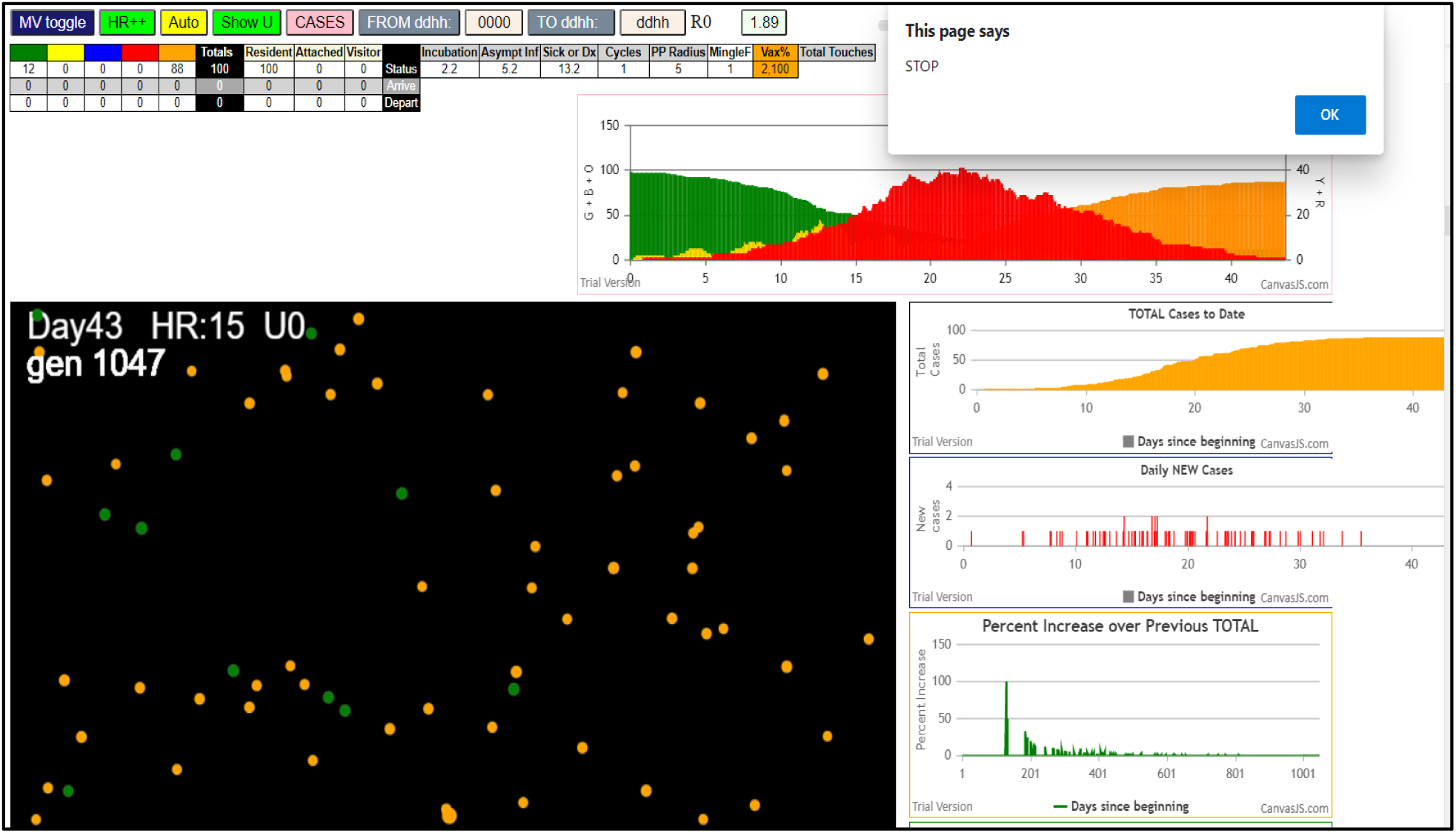
CovidSIMVL simulation of a self-extinguishing outbreak.

We initialize all simulations with a population of 100 persons in a finite space. Thus, density becomes an operative factor in determining spread. 99 agents are set initially as Susceptibles and one agent is set as an initial transmitter (Infective).

Three parameters are varied to change the intensity (rate of spread) of the underlying epidemic:

1. HazardRadius (HzR) – changes to this parameter capture degree of infectivity.
2. Symptomatic Days – this is keyed to the temporal dynamics of the infective agent.^10,11^ In CovidSIMVL, it reflects the number of effective symptomatic days. Thus, an upper bound is set by the duration of effective viral loads in the person. However, the actual value for any iteration within a simulation trial may be reduced to capture the effect of contact tracing or sustained physical separation.
3. Implementation of Vaccination Strategies – this is operationalized by changing the probabilities that proximity will produce transmission, with values conforming to the vaccination schedules – see *Implementation of Vaccine Strategies in Simulations*, below.

In other studies, we have also varied the MingleFactor parameter. This determines the amount of movement of agents within a finite space, subject to stochastic variation keyed to a Pareto distribution. For the trials in this paper, MingleFactor was set to 1 and did not vary. Together with the range of values assigned to HazardRadius, this assures sufficient mobility of agents so that the simulations do not regularly self-extinguish due to stochastic features of viral transmission, ***before*** the effect of the vaccination strategies has had a chance to show itself. Specifically, with the HazardRadius set to values used in the vaccination strategy trials (5, 4.5 or 4), and with no vaccination (Mode B), and no change in symptomatic days, most trials run close to 100% infected between 1000 and 2000 hours (24 generations in the simulation model per day), or 41 to 80 days.

As will be demonstrated, when vaccinations are applied, the trials may terminate by self-extinguishing because the transmitting agents have only a finite time period to reach a Susceptible and transmit (subject to probabilistically determined hurdles keyed to vaccination protection rates). Running out of transmitters means more survival for the population of vaccinated agents

### Simulation trials

Each trial is initialized with values on key parameters set to reflect vaccination strategies, degree of infectivity of agents, and contact tracing effectiveness. Trials proceed through iterations in which agents move through a space and encounter other agents. With each iteration, the state of an agent can change, given three factors: (a) their proximity to other agents; (b) the vaccination-schedule-dependent probability of infection when a critical proximity threshold has been crossed; and (c) biologically-determined progression of the infection within an agent.

Each trial terminates when there are no more Susceptibles, or no more Infective transmitters. The CovidSIMVL program generates a console.log for each transmission, in which each infective and infected agent is identified. As well, for each transmission there is a record in console.log of the probability used in that transmission to reflect degree of protection, if applicable.

### Implementation of Vaccination Strategies in Simulations

During the execution of a trial (simulation), if a transmission is feasible due to proximity of Infective or Susceptible agents, the program checks the current day in the simulation *vs* the day keyed to strategy milestones. CovidSIMVL uses a random number generator to decide whether to execute the transmission. Specifically, it compares the randomly assigned value to a criterion value keyed to the protection level conferred by a vaccine at a particular generation (time) within the trial. Thus, if the agents are in a situation where they have had both doses and 95% protection has been achieved, if the random number returns a number less than 0.95, the transmission does not occur, even if proximity would enable it to take place. If the number is greater than .95, a transmission will occur.

### Design

Trials are conducted with Mode1 and Mode2 strategies, under varying intensities of the underlying epidemic as set by the CovidSIMVL HazardRadius parameter which reflects differences in infectivity, and duration of infectivity for symptomatic agents (8 days or 4 days) to capture the effect of contact tracing. Intensity is varied within vaccination schedules to provide visibility into the potential for intensity to interact with the level and timing of protection conferred by a vaccine at different points in a vaccination schedule.

## RESULTS

### Summary

Each simulation trial runs until one of two possible conditions arise:

1. There are no more Susceptibles.
2. Although there are uninfected Susceptibles in the simulation, there are no more Infectives. In other words, the outbreak self-extinguishes before all Susceptibles are infected.

There are then two relevant summary outcomes. The first is the number of Susceptibles remaining when the trial self-extinguishes. The second is the number of generations required for the trial to self-extinguish. ***Table 1*** only contains information about remaining Susceptibles. The material in the ***APPENDIX*** also contains information about number of generations until self-extinction of the outbreak.

**Table 1.** Remaining Susceptibles (out of 100) at time of Self-Extinction of Simulated Epidemic.

|  | Baseline<br>No Vaccine | Mode1<br>1 Dose All<br>Trials | Mode1<br>“Worst<br>Case” | Mode2<br>2 Doses All<br>Trials | Mode 2<br>“Worst<br>Case” | Benefit:<br>2 Doses vs<br>None | Benefit:<br>2 Doses vs 1 |
| --- | --- | --- | --- | --- | --- | --- | --- |
| Pfizer | 4.1* | 7.6 | 0 | 50.4 | 20.0 | 46.3 | 42.8 |
| Moderna | 4.1 | 21.1 | 6 | 39.1 | 27.8 | 35.0 | 18.0 |
| Hybrid | 4.1 | 34 | 12 | 46.8 | 22.9 | 42.7 | 12.8 |

\*All figures represent mean number of remaining Susceptibles remaining at conclusion of a series of trials.

This first outcome is summarized in ***Table 1***. In this table, “All Trials” means the figures reflect the full set of 20 trials for Pfizer, 20 trials for Moderna, and 25 trials for Hybrid. The “Worst Case” figures are outcomes for only those trials where the parameters are set to reflect highest degree of infectivity (HazardRadius = 5) and no contact tracing.

Note that fractional values are obtained because the cell values are averages obtained across multiple runs of the different vaccine Modes for Pfizer, Moderna, or Hybrid, with different values for the HazardRadius parameter, and different numbers of Symptomatic Days to reflect the effects of contact tracing. These summary values in ***Table 1*** are based on results appearing in ***Tables 2, 3***, and ***4*** in the **APPENDIX**.

**Table 2.** Pfizer Trials.

| PFIZER TRIALS |  |  |  |  |  |
| --- | --- | --- | --- | --- | --- |
| Hazard Radius | RedDays | Mode | Terminate | End DD.HH | Survivors |
| 5 | 8 | B | 1047 | 43.15 | 3 |
| 5 | 8 | B | 1168 | 48.16 | 1 |
| 5 | 8 | 2,100 | 1090 | 45.10 | 15 |
| 5 | 8 | 2,100 | 1088 | 45.08 | 25 |
| 5 | 8 | 1,100 | 857 | 35.17 | 0 |
| 5 | 4 | B | 1141 | 47.13 | 3 |
| 5 | 4 | B | 1264 | 52.16 | 0 |
| 5 | 4 | B | 1188 | 49.23 | 6 |
| 5 | 4 | 2,100 | 885 | 36.21 | 30 |
| 5 | 4 | 2,100 | 726 | 30.06 | 48 |
| 5 | 4 | 1,100 | 1301 | 54.05 | 6 |
| 5 | 4 | 1,100 | 950 | 39.14 | 6 |
| 4.5 | 8 | 2,100 | 964 | 40.04 | 57 |
| 4.5 | 8 | 2,100 | 941 | 39.05 | 56 |
| 4.5 | 8 | 1,100 | 1182 | 49.06 | 3 |
| 4.5 | 4 | 2,100 | 729 | 30.09 | 71 |
| 4.5 | 4 | 2,100 | 773 | 32.05 | 63 |
| 4.5 | 4 | 1,100 | 1490 | 62.02 | 2 |
| 4 | 8 | B | 1138 | 47.10 | 1 |
| 4 | 8 | B | 2395 | 99.19 | 7 |
| 4 | 8 | B | 1639 | 68.07 | 2 |
| 4 | 8 | B | 1470 | 61.06 | 16 |
| 4 | 8 | 2,100 | 1025 | 42.17 | 40 |
| 4 | 8 | 2,100 | 1111 | 46.07 | 47 |
| 4 | 8 | 1,100 | 3366 | 140.06 | 8 |
| 4 | 4 | B | 1298 | 54.02 | 2 |
| 4 | 4 | 2,100 | 652 | 27.04 | 88 |
| 4 | 4 | 2,100 | 853 | 35.13 | 65 |
| 4 | 4 | 1,100 | 1704 | 71.00 | 12 |
| 4 | 4 | 1,100 | 1668 | 69.12 | 24 |

**Table 3.**
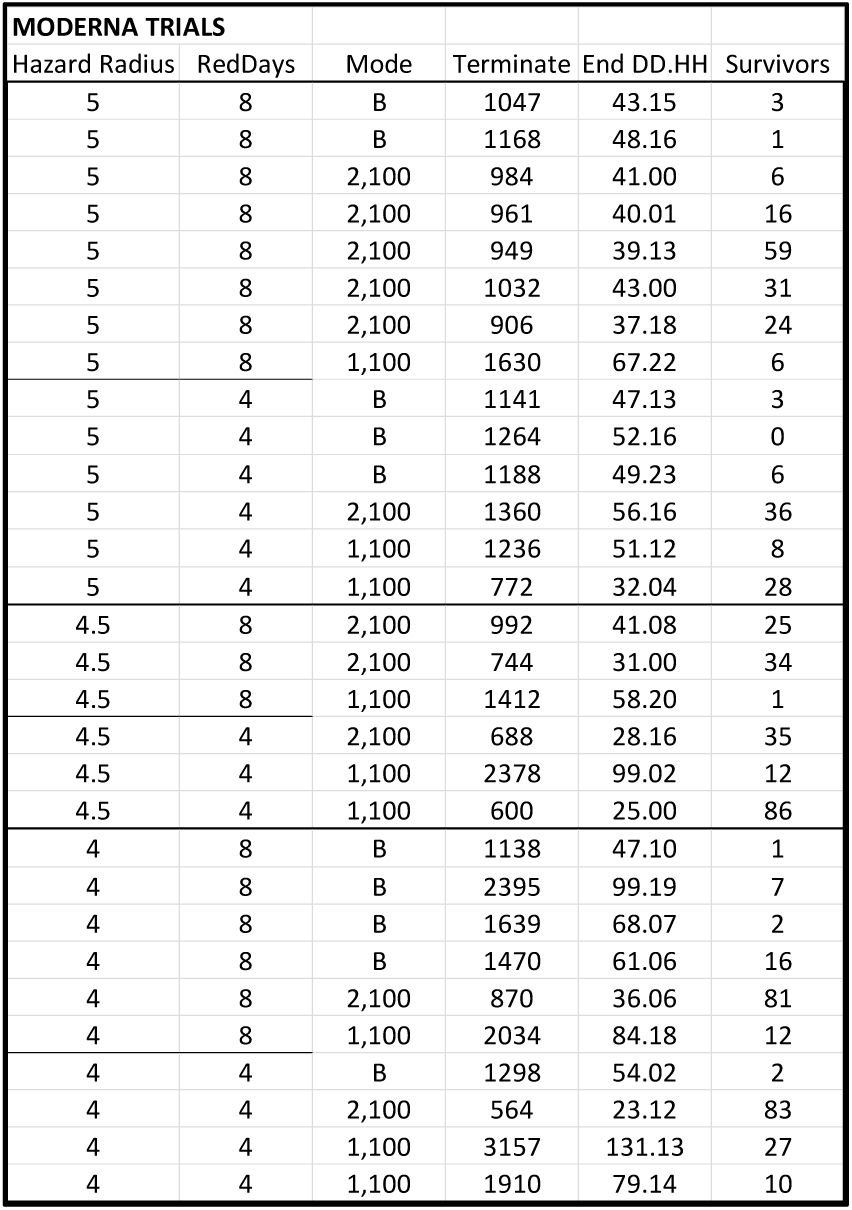
Moderna Trials.

**Table 4.**
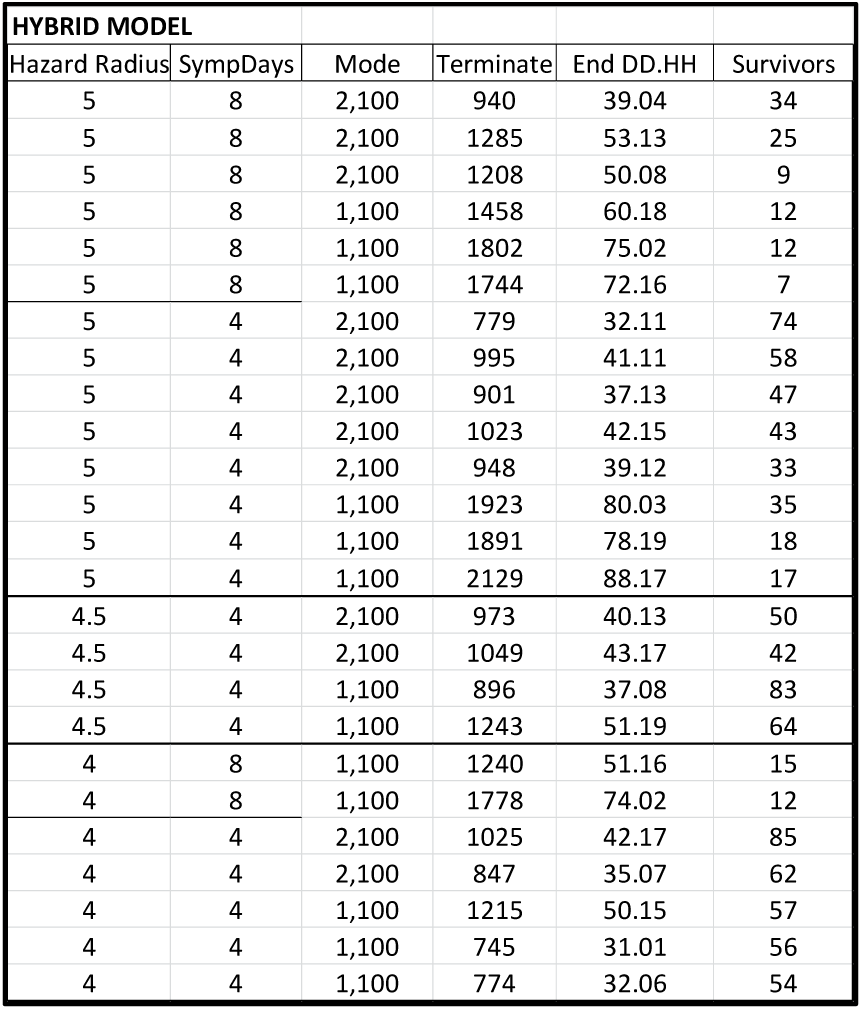
Hybrid Trials.

| HYBRID MODEL |  |  |  |  |  |
| --- | --- | --- | --- | --- | --- |
| Hazard Radius | SympDays | Mode | Terminate | End DD.HH | Survivors |
| 5 | 8 | 2,100 | 940 | 39.04 | 34 |
| 5 | 8 | 2,100 | 1285 | 53.13 | 25 |
| 5 | 8 | 2,100 | 1208 | 50.08 | 9 |
| 5 | 8 | 1,100 | 1458 | 60.18 | 12 |
| 5 | 8 | 1,100 | 1802 | 75.02 | 12 |
| 5 | 8 | 1,100 | 1744 | 72.16 | 7 |
| 5 | 4 | 2,100 | 779 | 32.11 | 74 |
| 5 | 4 | 2,100 | 995 | 41.11 | 58 |
| 5 | 4 | 2,100 | 901 | 37.13 | 47 |
| 5 | 4 | 2,100 | 1023 | 42.15 | 43 |
| 5 | 4 | 2,100 | 948 | 39.12 | 33 |
| 5 | 4 | 1,100 | 1923 | 80.03 | 35 |
| 5 | 4 | 1,100 | 1891 | 78.19 | 18 |
| 5 | 4 | 1,100 | 2129 | 88.17 | 17 |
| 4.5 | 4 | 2,100 | 973 | 40.13 | 50 |
| 4.5 | 4 | 2,100 | 1049 | 43.17 | 42 |
| 4.5 | 4 | 1,100 | 896 | 37.08 | 83 |
| 4.5 | 4 | 1,100 | 1243 | 51.19 | 64 |
| 4 | 8 | 1,100 | 1240 | 51.16 | 15 |
| 4 | 8 | 1,100 | 1778 | 74.02 | 12 |
| 4 | 4 | 2,100 | 1025 | 42.17 | 85 |
| 4 | 4 | 2,100 | 847 | 35.07 | 62 |
| 4 | 4 | 1,100 | 1215 | 50.15 | 57 |
| 4 | 4 | 1,100 | 745 | 31.01 | 56 |
| 4 | 4 | 1,100 | 774 | 32.06 | 54 |

This table indicates the following:

1. Under a No Vaccine condition, the parameters that govern dynamics of spread in CovidSIMVL have been set such that almost all agents becoming infected before the simulation self-terminates. This is indicated by the Baseline – No Vaccine value of 4.1, as in 4.1 remaining agents. In these ten No Vaccine trials, the number of remaining Susceptibles ranges between 0 and 16, where the variation reflects stochasticity of the viral transmission model built into CovidSIMVL
2. For all Mode1 trials (single dose) the dynamic reverts to the Baseline condition at 21 days for Pfizer, 28 days for Moderna, and 35 days for Hybrid. However, the number of remaining Susceptibles is greater than the number for the No Vaccination condition, reflecting protections conferred between the time when protections become operative in the model and the time when they terminate.
3. The slightly better result for Hybrid vs Moderna for a single dose and no second dose (Mode1), and far better result compared to Pfizer, reflects the assumption in the Hybrid model that the protection from the first dose will last 35 days for this Hybrid model, even though in Mode1, protection reverts to zero upon non-administration of the second dose. The Pfizer model assumes Dose 1 protection lasting only 21 days, and the Moderna model assumes 28 days protection from Dose 1. Though the Hybrid model assumes 75% protection (*vs* 80% for Moderna) upon administration of Dose 1, in these models the longer period of protection in the Hybrid model outweighs the benefit of a slightly higher rate of protection. Along these same lines, the shorter period of protection for Pfizer (21 days) along with the assumed 54% level of protection, yields the much lower number of remaining susceptibles at termination of the Mode1 trials.
4. For the single dose trials (Mode1), under “Worst Case” conditions (most virulent, no contact tracing) there were no remaining susceptibles at self-termination of the simulated outbreak for Pfizer, 6 remaining Susceptibles for Moderna and 12 for the Hybrid Model. These differences reflect the combined contribution of different degrees of protection following a single dose for Pfizer (54%) *vs* Moderna (80%) *vs* Hybrid (75%) and duration of protection from a single dose.
5. The summary Mode2 trials show the best result for Pfizer overall (i.e., averaging across degree of infectivity and contact tracing effects). Moderna shows the least benefit but it is critical to note that no contact tracing trials were completed for the Moderna series. Results for the Hybrid strategy are close to Pfizer. The superior performance of Pfizer, despite the lower level of protection between first and second dose compared to Moderna or Hybrid, is a reflection of the shorter prescribed time period between first and second dose, when protection goes up to 95%. However, in the case of a more virulent strain (Worst Case Scenario) this effect is counterbalanced. In these scenarios, Moderna performs best, reflecting the relatively higher level of protection from the time of the first dose.
6. Overall, the greatest benefit from completing the 2-dose regiment is found for Pfizer. This appears to reflect the benefits associated with achieving 95% protection a week in advance of Moderna and two weeks in advance of the Hybrid model.

## DISCUSSION

### Summary

A series 75 simulations were carried out to evaluate no vaccination vs full (two dose) vs partial (single dose) completion of different vaccination regimens. In addition to a schedule that conforms closely to material appearing in the Emergency Use Authorization documents from Pfizer and Moderna, a third regimen was included to provide some visibility into potential outcomes associated with a “Hybrid” strategy that entails a second dose being administered 35 days after the first (which falls outside the time boundaries for the Pfizer and Moderna stage 3 protocols).

The results illustrate some basic features of the outbreak dynamics when vaccinations are introduced as countervailing protective factors:

1. There is a delay between administration of the first dose and the protections afforded by that administration. The more virulent the strain of the virus, the less consequential is the time between first dose and second dose administration. If 100 persons receive a first dose but a large fraction are infected before that dose takes effect, the second dose will produce little benefit.
2. Conversely, a two-dose vaccine that produces the highest level of protection upon administration of the first dose will produce greatest benefit against a more virulent strain of the virus.
3. Viral spread reflects the joint contribution of several different factors, including effectiveness of vaccines, timing of administration, infectivity, and contact tracing. As shown in Moselle & Chang^12^ degree of movement of agents within a space of finite dimensions (MingleFactor parameter in CovidSIMVL) also alters the dynamics of spread.

### Methodological Note

The simulations involve “abrupt” transitions from one state of vulnerability to another. For example, the simulations confer no protection on agents until 14 days after inoculation, at which point agents acquire an average level of protection set to different values for Pfizer, Moderna or Hybrid strategies. As well, following specified intervals, protection drops to zero unless a second dose is administered.

These transitions are undoubtedly not correct and may introduce some measure of error in estimates of the relative impacts of the three vaccine schedules, or the impacts of not administering the second dose. Rather than trying to finesse out how the results might be impacted by this “abrupt transition” factor, we await further information on gradients to inject more gradualism and hence more realism into this set of simulations.

### Single Universe vs Multiverse version of CovidSIMVL – Potential Scope for Policy-Relevant Simulations

For all of the simulations covered in this document, agents were all lodged within a single space of finite dimensions (a CovidSIMVL “universe). However, the “real world” in which vaccination strategies have an impact is not homogeneous at a local level, and vaccination strategies are typically stratified (e.g., by age ranges) and sequenced in order to best manage risk on the basis of considerations of population-level transmission risk and on the basis of considerations of equity.^13,14,15^ In other words, mass action incidence models may rationalize the need for pan-societal protection and may be employed in generating estimates of level of protection *via* means of vaccination to achieve different degrees of herd immunity.^16,17,18^ However, “real-world” vaccination programs that stratify, target and sequence vaccine distribution on the basis of multiple criteria are not organized around what might be termed “mass population vaccination strategies”.

If the vaccination strategy is organized to reflect evaluation of risk associated with large segments of populations, then meta-population variants of equation-based models may provide requisite assessments of risk and relative benefits associated with different schedules.^19,20^ Where the intent of a strategy is to accommodate at some minimal level the real-world dynamics that govern local spread, models must come to terms with groups of people who are moving among different contexts that are distinguishable on the basis of dynamics governing transmission and hence are distinguishable on the basis of risk for transmission. ^21,22^ For example, a foundational “real-world” model that incorporated local dynamics in generating estimates of risk or benefits would consist of transmission chains that link contexts such as schools (for young children; for adolescents who have a transmission risk profile similar to adults), homes, places of recreation, places to acquire necessities of life, transportation facilities, and places of work. In the context of an outbreak, these chains are assembled into trees and the trees may then be meaningfully aggregated into forests of transmission trees, e.g., when a local population is exposed to a reservoir of infection that is triggering multiple outbreaks in settings that may be geographically separated or partially or completely overlapping.^23^ If the intent is to provide optimal protection at different stages in enacting a vaccination strategy, given issues related to supply and/or uptake, and if that optimization function entails targeting in enactment of vaccination strategies, and if the “last mile” in a vaccination strategy entails prioritized access to best manage local risk and achieve optimal aggregate level population outcomes, then what is required is a modeling tool that can nimbly execute simulated clinical trials in a heterogeneous space of transmission contexts. And, it should be noted, that tool cannot assume from the outset a reproduction rate (R0) when the objective is to use the simulation to discover what are local or aggregate reproduction rates. The “multiverse” version of CovidSIMVL, which treats reproduction rate as an emergent characteristic of transmission dynamics, provides a set of tools that enables such trials to be conducted. ^24^,25

The multiverse version of CovidSIMVL is tailored to addressing more complex policy-level issues related to vaccination strategies, such as priorities attached to reducing transmission *vs* protecting vulnerable persons or populations. For example, depending on vaccine supply and infectivity of prevailing strains, one option might be to vaccinate all of the patients in a long-term care facility. This would protect patients and prevent patient-to-staff transmission, and ensuing staff-to-community transmission.

However, another option to contemplate would entail vaccinating all family members and staff but not vaccinating patients (under conditions of limited vaccine supply) or providing a single dose to patients and both doses to family members and staff. The multiverse version of CovidSIMVL (like the single universe instance employed in this paper) can factor in infectivity and various time factors into simulations to determine which strategy, under which conditions provides most benefit conjointly to patients and to the community at large.

As well, given that strains of SARS-CoV-2 may vary in potential for transmission^26,27^ an important question is whether rising rates reflect different transmission dynamics in different contexts, or whether these rising rates reflect an inherent difference in infectivity as reflected in R0 value assumed by models that employ equations to reproduce measured rates. Real-world contexts of transmission do not correspond to the assumed world of homogeneous transmission in models that conform to the mass action incidence assumption.^28,29^ Because the multiverse version of CovidSIMVL is designed to reflect heterogeneous transmission, and enables individuals to display different rates of transmission in different contexts that locate agents more or less densely, possibly mingling at different rates, the tool can be used, at a minimum, to explore three potential classes of models related to the issue of potentially different degrees of infectivity:

1. Models where inherent potential for spread to take place is varied (*via* HazardRadius parameter), while the dynamic governing transmission with or across settings (density and movement of agents via the CovidSIMVL MingleFactor and other parameters) is held constant.
2. Models where potential for spread is held constant while dynamics determining actualization of that potential varies within or across settings (e.g., home vs recreational facility vs place of work).
3. Models in which both inherent potential for spread and dynamics govern actualization of that potential are varied.

Vaccination schedules can then be evaluated within these three groups of models to look at their contribution to protection in contexts where different factors are contributing to measured increases in rates in real-world settings.

### Next Steps

Work is underway to enable CovidSIMVL (written in Javascript) to be called as a function using the programming language R. This will enable more fully automated execution of multiple instances of trials set to the same parameter values, in order to look at distributions of output variables (e.g., remaining Susceptibles; generations until self-termination) and to determine what is a representative case *vs* an outlier for a given set of conditions. This will provide an alternative to a methodology that requires selection of a real-world historical dataset that is available from a documented outbreak and assuming that it encapsulates and reflects the core or essential dynamics of the viral transmission that gave rise to the data.

There are a large number of naturally occurring “experiments” taking place, as different countries or jurisdictions grapple with the challenges of vaccinating large numbers of persons with a limited vaccine supply while responding to a diverse array of interests and priorities. As capacity to run these vaccine-relevant simulations is built out *via* embedding of CovidSIMVL within a R analytical framework, new information will become available to set ranges on key parameters that have been subject to initial examination in this document. As well, different contributing/confounding factors will undoubtedly be identified which, when built into CovidSIMVL, will enable it to preserve or strengthen its relationship to real-world contexts, which was one of the driving motivations in pursuing an agent-based approach to COVID-19 modeling in the first place.

## Data Availability

The agent-based modeling tool (CovidSIMVL) and the files used to configure that tool for the simulations reported in this document are available on github (github.com/ecsendmail/MultiverseContagion)

### APPENDIX

The results reported in this manuscript are based on information appearing in the following tables. Column heading in ***Tables 2, 3*** and ***4*** may be understood as follows:

#### Hazard Radius

this value relates to infectivity. Higher values indicate greater likelihood for transmission to take place when agents are in sufficiently close proximity

#### RedDays

Symptomatic infectives in CovidSIMVL visualizations of infectious disease outbreaks are coloured red. The duration of infectivity is reflected in the value on RedDays. Note that some trials have a value of 8 days which is the value used to simulate no contact tracing. RedDays = 4 is intended to reflect outbreaks where contact tracing is effective in shortening the number of days in which persons can infect others.

#### Mode

this indicates whether the simulation is one in which agents are not vaccination (Mode = B = baseline), or Mode = 1,100 (Single does of a 2-dose vaccine, 100 agents at start), or Mode = 2,100 (Two doses of vaccine, 100 agents at start)

#### Terminate

number of generations before the simulation terminates. This occurs when there are no more Susceptibles or when there are no more agents who are infective. There are 24 generations in a day.

#### End DD.HH

this is a translation of the number of generations into Days and Hours.

#### Survivors

this is the number of Susceptibles remaining at the time that the simulation terminates.

